# Constructing Probabilistic Human Mobility Patterns for Estimating Total Exposures and Pathogen Transmission Using Aggregate Mobile Phone Data

**DOI:** 10.1101/2024.11.26.24317970

**Authors:** Julio C. Facelli, Ram Gouripeddi

## Abstract

While phone location data is widely collected by providers and shared under special agreements with data aggregators, it is not available for research or routine surveillance purposes. Moreover, raw phone data may expose personal travel patterns which would not be ethically or lawful to use. One large data aggregator, Advan (https://advanresearch.com/) provides aggregate traffic patterns at weekly and monthly temporal resolution for many points of interest. This aggregate data is mostly used for commercial applications, but we argue that it can be also used to develop a model of probabilistic human mobility patterns at weekly or monthly resolution. Here we provide a stochastic algorithm that using the aggregate data provides realistic human mobility patterns at the intended temporal resolution. In the proposed model we associate a “person” with both a unique census block as the residence and another census block as the workplace. These human mobility patterns are dynamic and can be used in exposure models and in epidemiological models of epidemic spreads. Because these models do not assume any exposure or pathogen transmission mechanisms, they can be coupled to any external exposure fields or transmission model of infectious disease. Here we show how they can used to build a probabilistic model of exposure for an arbitrary exposure filed E(X, t) and a SIR contagion model. This probabilistic model can be developed at monthly or weekly time resolutions depending on the data source available and the specific programmatic needs.

## Introduction

Estimating human exposure to airborne and other broadly distributed pollutants as well as developing realistic models of pathogen transmission presents a significant public health challenge (Dons et al., 2011; Dons et al., 2014). Because humans are mobile and inhabit a variety of microenvironments, it is insufficient to model only the spatiotemporal distribution of pollutants or pathogens. Even for a geographically homogeneous distribution of pollutants, different individuals experience different levels of total individual exposure depending on their activity patterns (Dias & Tchepel, 2018; Mennis et al., 2018; Qian et al., 2017; Réquia Júnior et al., 2015). This is also true for mathematical models used to understand the spread of infectious diseases as we demonstrated in our work on SARS-CoV-2 (Lund et al., 2017; Soares et al., 2014; Wang et al., 2021).

Modeling is the only viable scientific approach to develop effective strategies that can minimize these undesirable side effects and show the effectiveness of interventions (Gates, 2015, 2018) to mitigate the effects of environmental exposure and pathogen contagion. But modeling should be informed by human activity patterns as different humans will come in contact with infected individuals or be exposed to different substances depending upon their daily activities. To the authors’ knowledge, very few models have incorporated empirical information about human activity patterns in their simulations. We have shown that human activity patterns, as recorded by the American Time Use Survey (ATUS) (Statistics, 2016), can be classified and used to simulate human activity in the context of estimating personal exposure to PM_2.5_ (Lund et al., 2020). Based on this classification of human activities, we have developed the Spatial Temporal Human Activity Model, or STHAM (Lund et al., 2020) model to generate and characterize travel trajectories and activity patterns (Bellemans et al., 2010; Bhat et al., 2004; Bradley et al., 2010), and to integrate exposure-agent geographical distributions to estimate total human exposure in large populations. The fundamental weakness of the work mentioned above is that the simulations are based on static human activity patterns, which may change dramatically upon the start of a pandemic or an environmental exposure exacerbation due government regulations, personal decisions and public health warnings. Furthermore, a large amount of pollutant emissions in urban environments result directly from human activity (Abi-Esber & El-Fadel, 2012; Apte et al., 2017). Modeling human activities could also be used in estimating pollution distributions directly from mobile sources, like automobile emissions (Greco et al., 2007; Tainio et al., 2010).

Modern technologies such cell phones, traffic measurements, surveillance cameras can provide accurate and timely information of human movement. This information is routinely collected by service providers and security agencies (Goldenholz et al., 2018; Yabe et al., 2024), but not available to public health authorities or researchers due to privacy and security concerns. Therefore, any successful and widely applicable model of individual human exposure will require a realistic estimation of the sequences of the activities that human agents perform, along with their locations and context derived from aggregate data. Therefore, methods are needed to model human mobility based on available information from aggregate data sets. One large data aggregator, Advan https://advanresearch.com/) provides aggregate traffic patterns at weekly and monthly temporal resolutions for many points of interest (POI). This aggregate data is mostly used for commercial applications, but we argue that it can be also used to develop a model of probabilistic human mobility patterns at weekly and monthly resolution.

Here we present a stochastic algorithm that using this type aggregate data can provide realistic human mobility patterns at intended temporal resolutions. In the proposed model we associate a “person” to both a unique census block of their residence and another census block of their workplace. These human mobility patterns are dynamic and can be used in exposure models and in epidemiological models of epidemic spreads. Because these models do not assume any exposure or transmission mechanisms, they can be coupled to any external exposure fields or transmission models of infectious disease. Here we show how the proposed model can be used to build a probabilistic model of exposure for an arbitrary exposure field E(X, t), where E is the strength of the exposure at time t in the location X, and a SIR contagion model (Amador, 2014). While we use the data structure available in the Advan data sets, the approach is generalizable to any similar aggregate data set derived from phone tracking data.

### Model Equations

We developed a model in which we associate a “person” with both a unique census block as the residence HCBGS and the workplace by another census block indicated as WCBGS. When no information exists about the workplace location of a “person” this can inferred by the proximity and number of “persons” not visiting POIs in the proximity of the “person’s” WCBGS during working hours (weekly) or working days (monthly). We structure the data as a series of epochs or periods such as:

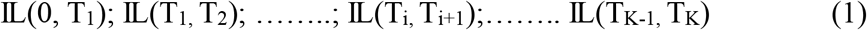

where IL(T_i,_ T_j_) could be HCGBS_α_ (T_i,_ T_i+1_), HCGBS_β_ (T_i,_ T_i+1_) or POI_γ_ (T_i,_ T_i+1_) and K is the number of epochs considered in the model. Therefore, for each epoch defined by the interval [T_i,_ T_i+1_]

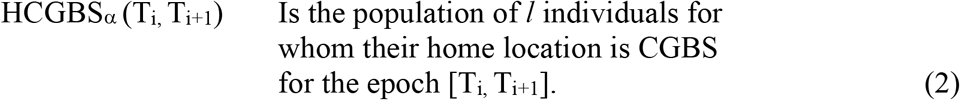

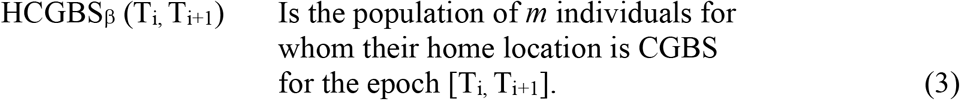

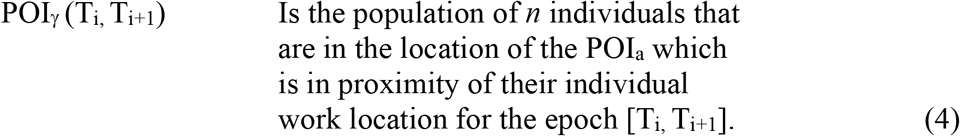

At any giving time, *l* + *m* + *n* = N, where N is the total number of individuals in the model. We assume that the distribution of visitors at a POI which is POI_γ_ (T_i,_ T_i+1_) for each HCBGS_α_ and WCBGS_β_, is a power distribution from the distance between the IL (T_i,_ T_i+1_), IL = HCBGS_α_ or WHBGS_β_ and the POI_γ_ (T_i,_ T_i+1_). Therefore, it is possible to define a cutoff distance that allows to associate each POI_a_ (T_i,_ T_i+1_) with a small set, perhaps ∼10-20, HCBGS and WCBGS in its proximity and assume that only these POIs are frequented by “persons” living or working at the HCBGS_α_ or WHBGS_β_, respectively.

So, a “person” who lives in certain HCBGS_α_ or works in a WCBGS_β_ can be associated with a set of Ω POIs in proximity to the HCBGS_α_ and Λ POIs in proximity to WCBGS_β_. Where these populations are given by:

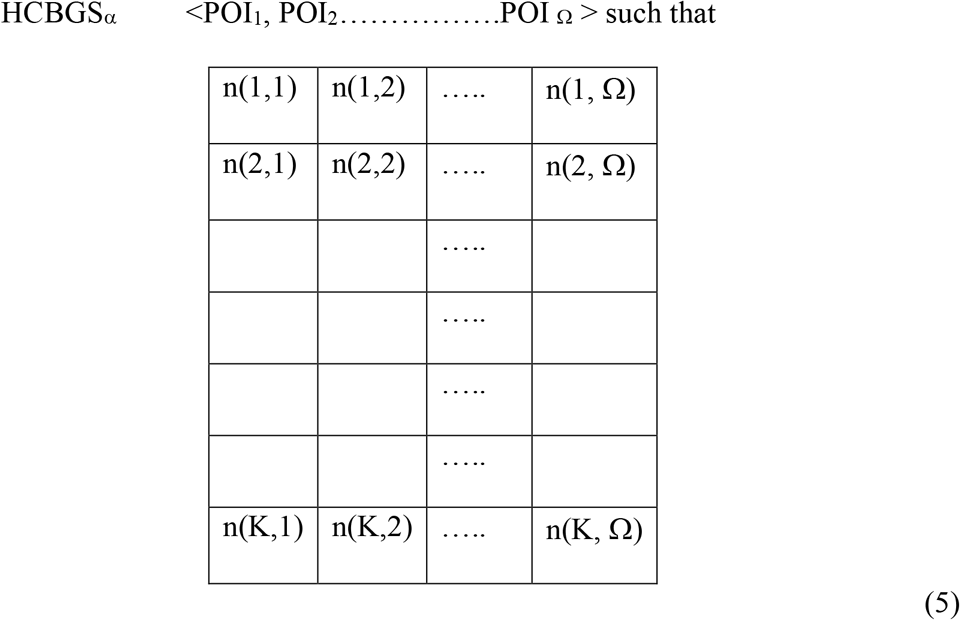

where n(i, _γ_) is the number of visitors from HCBGS_α_ to POI_γ_ in epoch i; where i ranges from 1 to K, and K = 168 or 30, for the weekly (hourly resolution) and monthly (daily resolution) aggregates, respectively.

Similarly, we have a matrix describing the visitors to its proximity POIs from a workplace WCBGS_β_, were n(i, _γ_) is the number of visitors from WCBGS_β_ to POI_γ_ in epoch i, where i ranges from 1 to K,

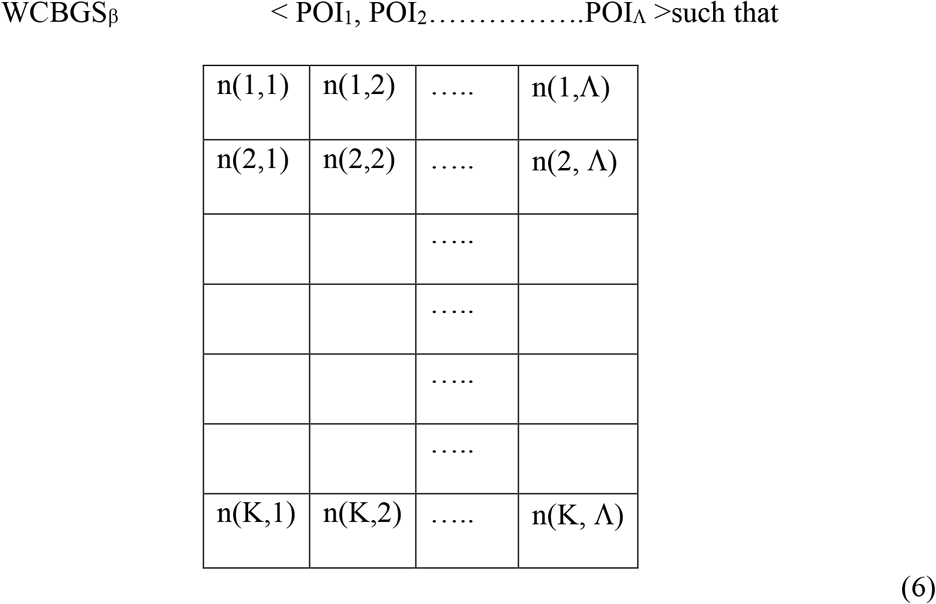

Assuming similar behaviors for all the “persons” living in a HCBGS_α_ we can approximate the probability of a “person” in HCBGS_α_ visiting a POI (i) at time j will be

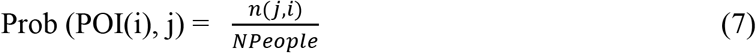

where *NPeople* is the number of “persons” living in HCBGS_α_. If we do not have information about of the number of “persons” that reside in HCBGS_α_ that work, we can infer this number or more precisely the fraction, ω, of workers residing in HCBGS as:

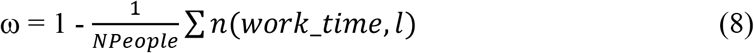

where n(*work_time*, l) is the number of “persons” visiting the POI (l) during the *work_time* times which correspond to the expected working hours during the day for the weekly aggregates and expected working days for the monthly aggregates. From other sources (McNeill et al., 2017; Mitra & Saphores, 2019; Yang et al., 2014) we can get the average commuting distances for workers residing in HCBGS_α_ and with this distance we can infer a set of likely workplaces, WCBGS_β_, from which we can randomly choose one that will be associated with the “person” residing in HCBGS_α_ with a probability ω that the person is working WCBGS_β_. If the workplace is known the derivations above are not necessary and a “person” residing in HCBGS can be associated with a unique WCBGS _β_, with a probability ω = 1.

### Exposure Equations

Using the definitions given above we demonstrate their use in building a probabilistic model of exposure for an arbitrary exposure field E(X, t), where E is the strength of the exposure at time t in the location X, or a contagion model (later). The probabilistic model can be developed for monthly or weekly time resolutions depending on the data source used as discussed above and the specific programmatic needs of the model. Models with higher resolution, i.e., daily, could also be implemented following the same framework presented here if the data would be available.

For an individual living HCBGS_α_, and having a probability ω of working at WCBGS_β_, at time t the individual will be exposed to the following concentrations of a pollutant under consideration:

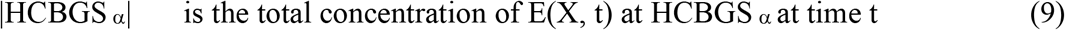

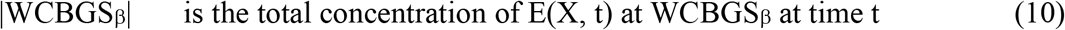

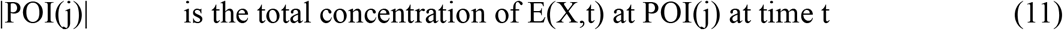

This individual will be exposed to these concentrations of E(X,t) for a time τ (1 hour for weekly resolution or 1 day for monthly resolution, therefore the total exposure at time k will be:

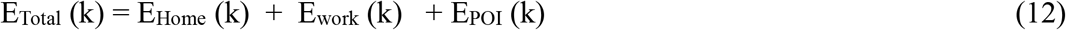

which are defined as:

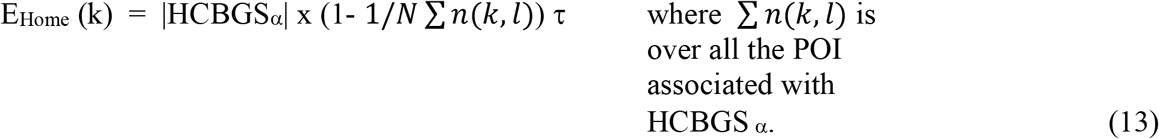

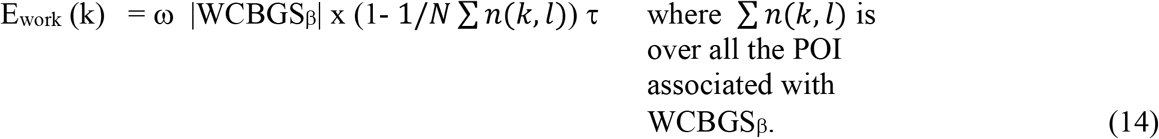

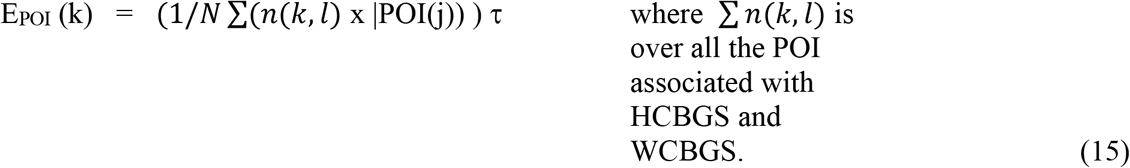

The total exposure can be calculated as

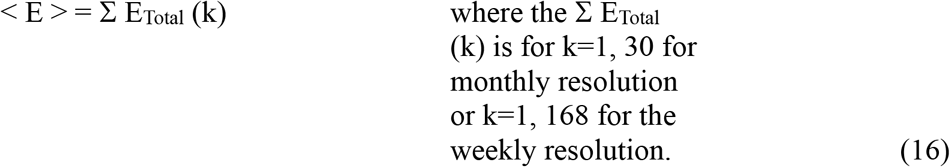

### Transmission Rates Using an SIR model

In this section we demonstrate how our human mobility model can be used to develop an SIR contagion model (Amador, 2014). For simplicity we show how the human mobility model can be used in the formulation this simple contagion model, the SIR model, but clearly there is nothing in the development that would preclude using more complex models by replacing the linear equations of the SIR model, Eqs. 26 and 27 below, for those of more complex models.

We will use the following definitions:

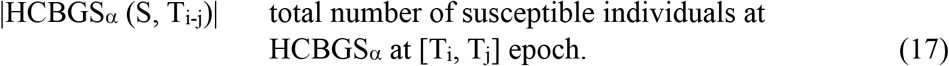

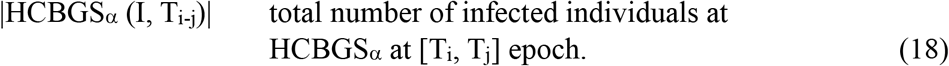

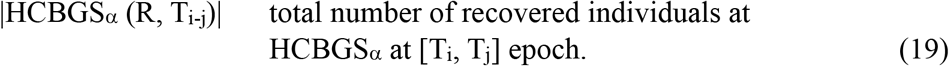

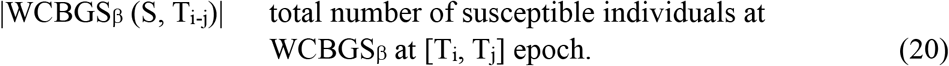

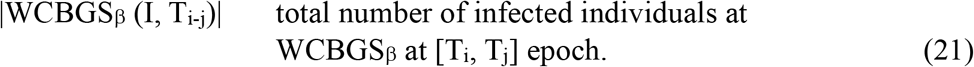

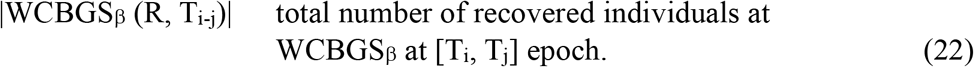

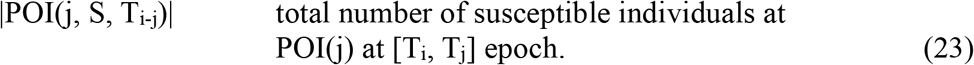

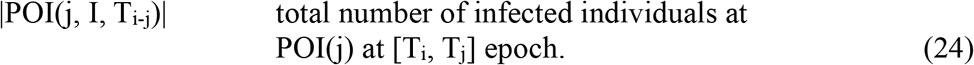

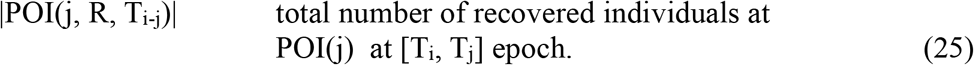

We define IL(S, T_i_), IL(I, T_i_), and IL(R, T_i_) as the generic designation of the total number of S, I and R individuals at any of these locations. Next, we compartmentalize these individuals, and formulate the corresponding first order differential equations to evolve these populations according to a SIR model for each of the location considered here - residential, and work census tracks, and POI in which we argue different “persons” are more likely to be in contact with each other than those living or working at distance locations.

Assuming that all infected individuals are able to transmit the pathogen, an individual will have a probability Ω of transition between, S (susceptible) to I (infected) and a probability Φ to transition between I (infected) to R (recovered) during a time period τ (1 hour for weekly resolution or 1 day for monthly resolution, therefore the total transition can be calculated during each epoch [T_i_, T_j_]. For each epoch and for each physical compartment (i.e. location), IL(S, T_i_), IL(I, T_i_), and IL(R, T_i_), we will have for t such that T_i_ < t < T_j_ and considering that the initial populations at T_i_ is IL(S, T_i_); IL(I, T_i_); IL(R, T_i_), which are the initial populations of susceptible, infected and recovered population at the beginning of the epoch, at T_i_ and where IL corresponds to HCBGS, WCBGS or POI populations and assuming that there is not mixing of the populations, a conditions that is reasonable for the hour resolutions data, but perhaps difficult to defend for the weekly one. According to the SIR model (Amador, 2014) the populations will evolve accordingly to the following first order equations:

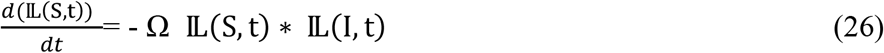

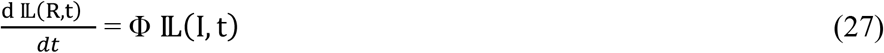

These differential equations are used to calculate the populations of susceptible, infected, and recovered individuals at the end of the epoch, obtaining IL(S, T_j_); IL(I, T_j_); IL(R, T_j_), for all ILs considered here. We should calculate the evolution of the populations for all possible HCGBS, WCGBS and POIs.

### Evolution of the Population Between Epochs

The information available from the observation logs gives us the distribution of the population for each epoch but does not provide any information on which individuals will change location among the different compartments. Here we present two scenarios for the transition of individuals between epochs, [T_i_, T_j_] to [T_j_, T_j+1_]. In the first scenario we assume locality, i.e., the probability of the transition between compartments IL(T_i,_ T_j_) to IL(T_j,_ T_j+1_) is proportional to the physical distance between these compartments. For the second case we assume that there is a global random redistribution. It should be noted that these are just two possible examples, and perhaps, represent two asymptotic movement distribution or redistribution, but any distribution functional that preserves the total number of susceptible, infected, and recovered individuals will be valid. Here we present these two extreme cases as no information about transitions is available in our data, but more realistic functions could be used if experimental evidence is available to justify for them. For the random distribution of S, I and R individuals the new IL(T_j,_ T_j+1_) will be associated with l susceptible individuals S_Ti_, m infected I_Ti_ and n recovered ones R_Ti_, where

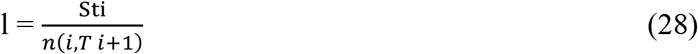

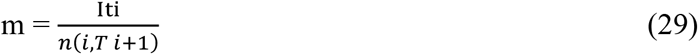

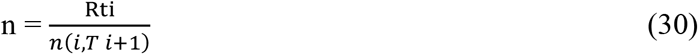

where n(i, T_j+1_) is the number of individuals in the corresponding IL(T_j,_ T_j+1_), where IL(T_j,_ T_j+1_) could be HCGBS_α_(T_j,_ T_j+1_), HCGBS_β_(T_j,_ T_j+1_) or POI_n_(T_j,_ T_j+1_). Alternative if we use the locality approximation, in which we assume that the transition probabilities between IL(T_i,_ T_j_) and IL’(T_j,_ T_j+1_), where IL’(T_j,_ T_j+1_) could be HCGBS(T_j,_ T_j+1_), HCGBS(T_j,_ T_j+1_) or POI_n_(T_j,_ T_j+1_) is proportional to the distance between their locations.

Let us define:

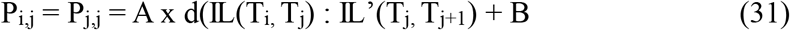

where d(IL(T_i,_ T_j_) : IL(T_j,_ T_j+1_) is the physical distance between IL(T_i,_ T_j_) and IL(T_j,_ T_j+1_).

We will have to execute a doble loop over all possible IL and IL’ calculating the transition probabilities such that in IL(T_i,_ T_j_) there are l susceptible, m infected and n recover individuals l x P_i, j_; m x P_i, j_ and n x P_i, j_ individuals will transfer from IL(T_i,_ T_j_) to IL’(T_j,_ T_j+1_). Note that the doble loop is necessary to consider the transfer in the opposite direction. In general, the populations at the beginning of the new epoch IL’(T_j,_ T_j+1_) will be α susceptible; β infected and γ recovered individuals, where α, β and γ are defined as:

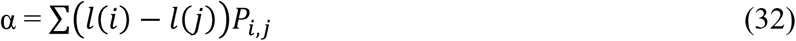

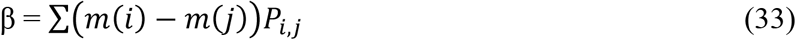

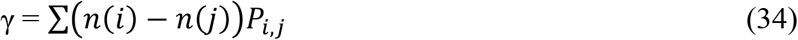

where the sums are over all possible pairs of IL(T_i,_ T_j_) and IL(T_j,_ T_j+1_) compartments.

Using the equations formulated for the exposure to an external field, we can also calculate the probability of infection, PI of individuals living in HCBGS_α_ with a ω probability of working at WCBGS_β_ and frequenting a POI by replacing E(X, t) by the number of infected individuals at location X, times the contagion parameter Ω times the time spend in this local for a given epoch.

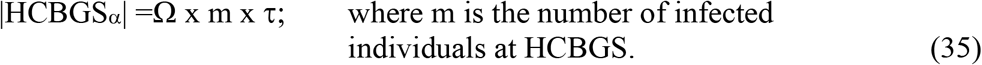

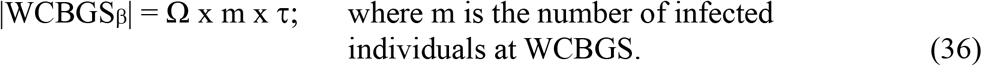

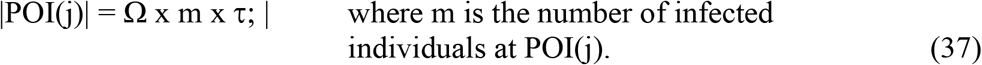

which are defined as

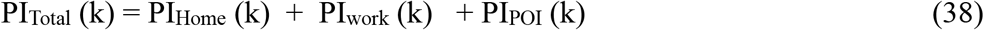

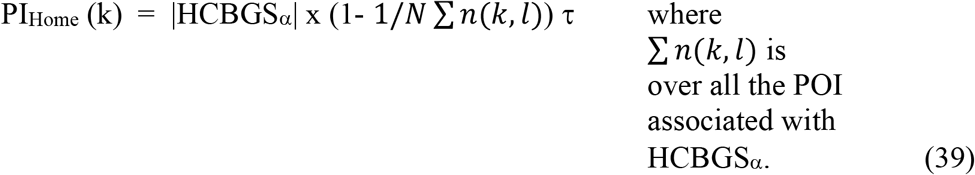

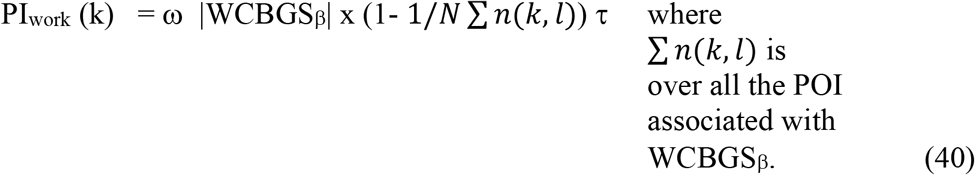

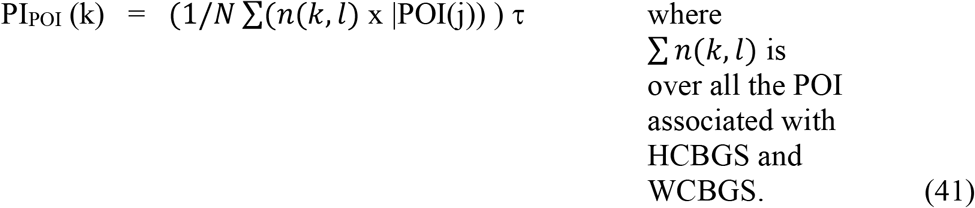

The total probability of infection, PI can finally be calculated as

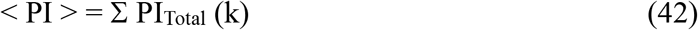

where the Σ E_Total_ (k) is for k=1, 30 for monthly resolution or k=1, 168 for the weekly resolution

## Discussion and Conclusions

We presented a stochastic model that using aggregate mobile phone data can provide an empirical and consistent human mobility pattern. While we used the Adavan data model of aggregate phone location data to develop the model, it can be adapted to any other phone or camera aggregated human mobility data. The temporal resolution used here was hourly and daily, based on availability in the Adavan data, but these are not intrinsic limitations in the human mobility model itself. We have demonstrated how this model, which can serve a primary model in any meta-modeling framework by using it to develop a full exposure model for any arbitrary exposure field, and a simple SIR model for infection transmission. These should be seen as demonstration of the possibilities of the model, but certainly the model is able to accommodate more complex models of exposure or transmission.

Limitations of this work include that we have used arbitrary assumption to associate house and work locations with the POI of interest, while this is an obvious limitation any further information on this may move the modeling close to identifiable human movements that may collide with privacy and security concerns. The same argument can be used to justify the use of census block as the higher spatial resolution. The other important limitation of the model, which was exposed in the development of the SIR infection model, is that the data set does not provide any information on how the population evolves (moves between compartments) between the end and start of consecutive epochs. While more granular information about this may be useful, it also will increase the risk of re-identification.

## Data Availability

No data is available because this is atheoretical paper. This also true for questions #2 and #3, we were forced to answer YES, but really should be no applicable.

## Acknowledgements

The research reported here was supported in part by the National Center for Advancing Translational Sciences of the National Institutes of Health under Award Number UM1TR004409. The content is solely the responsibility of the authors and does not necessarily represent the official views of the National Institutes of Health.

